# Accuracy of Medical Billing Data Against the Electronic Health Record in the Measurement of Colorectal Cancer Screening Rates

**DOI:** 10.1101/19004598

**Authors:** Vivek A. Rudrapatna, Benjamin S. Glicksberg, Patrick Avila, Emily Harding-Theobald, Connie Wang, Atul J. Butte

**Affiliations:** Division of Gastroenterology, University of California, San Francisco, CA, 94158; Bakar Computational Health Sciences Institute, University of California, 550 16th Street, San Francisco, CA, 94158; Department of Pediatrics, University of California, San Francisco, CA, 94158; Center for Data-Driven Insights and Innovation, University of California Health, Oakland, CA, 94607

**Author notes:** Corresponding Author: Atul J. Butte. 550 16th Street, 4th Floor Box 0110, San Francisco, CA 94158-2549. These authors contributed equally to this work (Vivek A. Rudrapatna, Benjamin S. Glicksberg). **Contributions:** VAR and BSG conceived the project, performed data extraction and analysis, and drafted this manuscript. VAR, PA, EHT, and CW performed the chart review. AJB supervised the project and critically edited this manuscript. **Ethics:** Approved by the University of California, San Francisco Institutional Review Board (#18-25166).

**Keywords:** Electronic Health Records, Colorectal Cancer Screening, Clinical Informatics

## Abstract

**Objective:** Administrative healthcare data are an attractive source of secondary analysis because of their potential to answer population-health questions. Although these datasets have known susceptibilities to biases, the degree to which they can distort measurements like cancer screening rates are not widely appreciated, nor are their causes and possible solutions.

**Methods:** Using a billing code database derived from our institution’s electronic health records (EHR), we estimated the colorectal cancer screening rate of average-risk patients aged 50-74 seen in primary care or gastroenterology clinic in 2016-2017. 200 records (150 unscreened, 50 screened) were sampled to quantify the accuracy against manual review.

**Results:** Out of 4,611 patients, an analysis of billing data suggested a 61% screening rate. Manual review revealed a positive predictive value of 96% (86-100%), negative predictive value of 21% (15-29%), and a corrected screening rate of 85% (81-90%). Most false negatives occurred due to exams performed outside the scope of the database – both within and outside of our institution – but 21% of false negatives fell within the database’s scope. False positives occurred due to incomplete exams and inadequate bowel preparation. Reasons for screening failure include ordered but incomplete exams (48%), lack of or incorrect documentation by primary care (29%) including incorrect screening intervals (13%), and patients declining screening (13%).

**Conclusions:** Although analytics on administrative data are commonly ‘validated’ by comparison to independent datasets, comparing our naïve estimate to the CDC estimate (∼60%) would have been misleading. Therefore, regular data audits using the complete EHR are critical to improve screening rates and measure improvement.

**Study Highlights:** *WHAT IS KNOWN:*

- Medical billing data might be useful for measuring colon cancer screening rates but are bias-prone and difficult to validate
- The degree to which these biases may skew the results of simple population-level analytics is not widely appreciated, nor are their causes and possible solutions.

*WHAT IS NEW HERE:*

- Billing data from the health record does not accurately capture unscreened patients. Some reasons were predictable (screening outside the system or prior to software implementation) but others were not.
- The common practice of external validation would have been falsely reassuring for these data. The naïve estimate of screening rates matches the CDC estimate (61%); the true rate was 85%.
- Periodic data audits using the full EHR is critical to continue to improve screening rates and monitor improvements accurately and at scale.

## Introduction

Colorectal cancer (CRC) screening is a high priority for public health in the US and abroad. Although CRC remains the second leading cause of cancer-related death in the US^1^, screening via modalities such as colonoscopy have the potential to reduce the mortality rate by 60% or more^2^. Despite its potential for such impact, screening uptake as estimated by the Centers for Disease Control (CDC) has remained stagnant at 60% for at least a decade.^3,4^ These findings have prompted multiple calls for action such as the 80% by 2018 campaign led by the National Colorectal Cancer Roundtable.

The traditional benchmark for measuring CRC screening rates in the US has been the National Health Interview Survey – an annual survey of the civilian and non-institutionalized population. Although these data are considered the gold standard, they suffer from a number of shortcomings including low participation rates (55%)^4^, recall bias, lack of confirmation with the medical record, uneven health literacy, and social desirability bias^5^.

An alternative source that avoids many of the aforementioned pitfalls is administrative healthcare data. Although these data were originally collected to support operations and financial objectives, they could potentially be useful for many other purposes: tracking the effectiveness of screening outreach measures, providing clinical decision support, and rewarding providers and health systems for value-based care^6^.

However, precisely because these data were originally assembled for other reasons, they are prone to measurement bias^7^. More concerning, many large structured datasets such as those derived from medical claims can be difficult to validate, in part due to disconnection from the underlying medical context. Therefore, even though transparent and repeated benchmarking is a critical step for any valid data repurposing endeavor, this is rarely done.

Although it can be difficult to benchmark the accuracy of claims data from payor databases, billing data derived from the EHR may represent a good proxy for two reasons: 1) much of claims data are derived from bills generated by EHR software over the course of clinical operations, and 2) algorithms based on these data may be validated against the full clinical context captured in the EHR.

In this study, we attempt to answer the question: how accurately do medical billing data capture the colorectal cancer screening rates within a healthcare system? Here, we perform an informatics-based estimation of the period prevalent screening rate using billing data derived from the EHR. We then review a random sample of charts in order to identify the reasons for algorithmic misclassification and missed screening. We conclude by proposing strategies to enhance future clinical informatics efforts and improve the primary prevention of colorectal cancer.

## Methods

### Clinical Data

EHR data was extracted from the University of California, San Francisco (UCSF) Epic system using Clarity and Caboodle tools^8^. To model the kind of data available in typical payor claims databases, we extracted the following structured fields: age, gender, ‘Alive’ status, race, primary language, ethnicity, insurance, department, diagnosis code, procedure code, and encounter date.

Prior to being used for this study, the data was de-identified to comply with the US Department of Health and Human Services ‘Safe Harbor’ guidance. Temporal imprecision was introduced into the dataset via a random negative offset (0-364 days).

### Study Population

We included patients aged 50-74 who had at least two primary care visits, two gastroenterology clinic visits, or one of each between January 2017 and December 2018 (see Figure 1). We excluded charts bearing the following International Classification of Disease, Tenth Revision, Clinical Modification (ICD-10-CM) codes reflecting an elevated risk of colorectal cancer: Family history of colon polyps (Z83.71, Z83.79), Family history colon cancer (Z80.0, Z80.9), Personal history of colon polyps (Z86.010, K63.5, D12, K63.5), Personal history of colon cancer (C18-C21), Hereditary Non-Polyposis Colorectal Cancer/Lynch Syndrome (Z15.09 Z14.8, Z80.0, Z84.81), Familial Adenomatous Polyposis (D12.6, Z14.8), Juvenile Polyposis Syndrome (D12.6), Peutz-Jehgers Syndrome (Q85.8, L81), and Inflammatory Bowel Disease (K50, K51). We curated records corresponding to code Z98.89; patients who were annotated as having either a history of prior lower endoscopy or colectomy and lacked an order for a screening exam were excluded.

**Figure 1:**
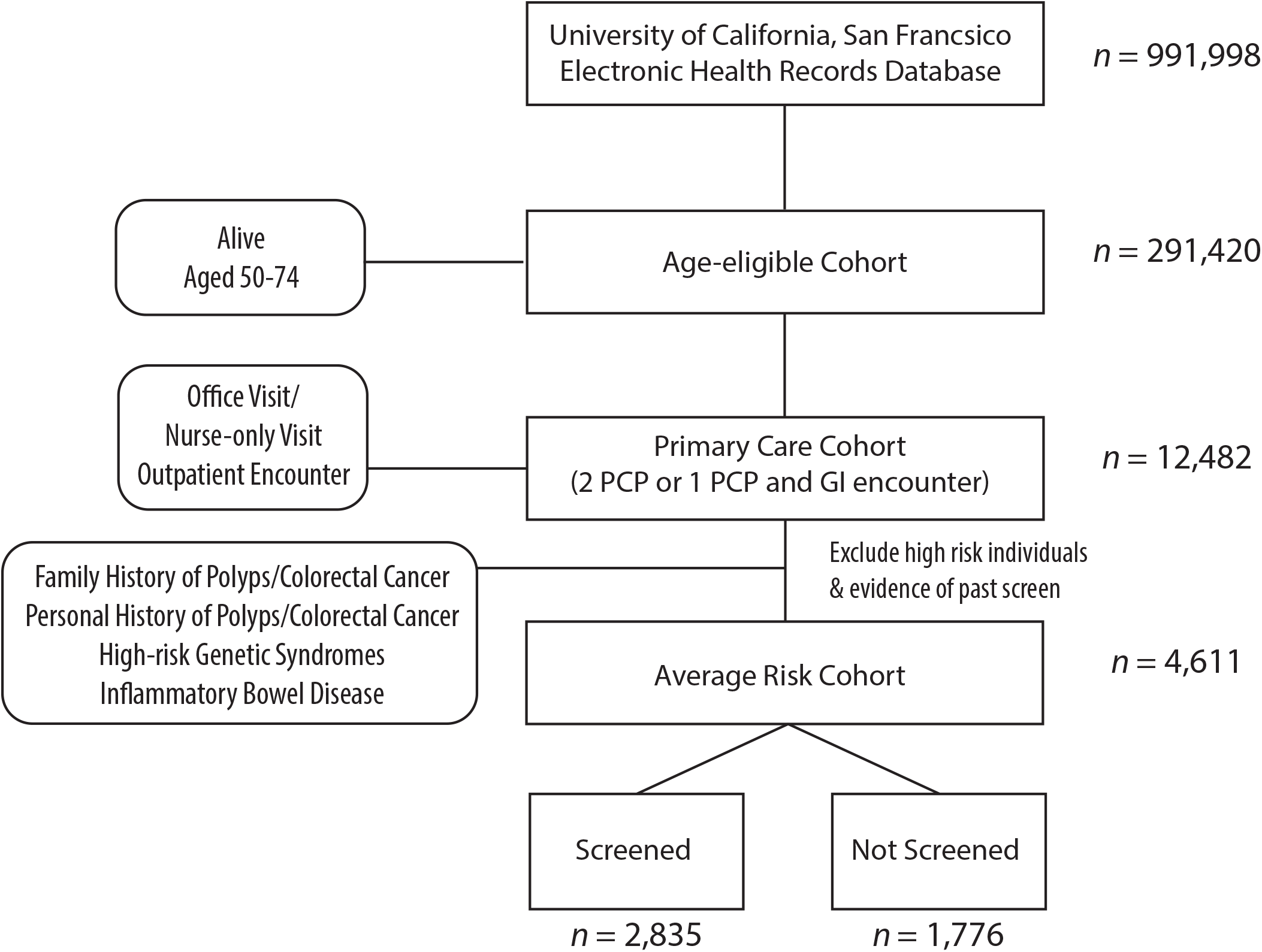
Cohort Selection Schematic.

### Classification Algorithm

We identified charts with a prior history of lower endoscopy using Current Procedural Terminology (CPT) codes (see Supplemental Methods). Additionally, we used regular expression-based string matching to identify billed for procedures corresponding to Colonography-protocoled Computed Tomography (CT Colonography), Double Contrast Barium Enema, and Fecal Immunochemical Test. Capsule colonoscopies, guaiac-based stool testing, and fecal DNA tests are not performed at our facility.

We used the following schedule to determine the presence or absence of a qualifying screening exam: Colonoscopy within the prior 10 years, Sigmoidoscopy within the prior five years, Fecal Immunochemical Test (FIT) in 2016, CT Colonography within the last five years, Double Contrast Barium Enema within the last five years. Patients were classified as screened if they had been screened according to this schedule as of March 2018.

### Database Querying and Analysis

All queries required several rounds of iterative refinement done in close collaboration between the clinical and bioinformatics teams. Identification and verification of CPT codes were performed in close consultation with gastroenterology billing specialists. ICD-10 codes were selected by manual review. Encounter names corresponding to primary care visits were identified by discussion with primary care physicians. Data extraction was performed using MySQL (version 5.6.10). Further refinement and analysis was performed in the *R* programming environment^9^ (version 3.4.1) using the *RMySQL*^10^ and *data*.*table*^11^ packages. Agresti-Coull binomial confidence intervals^12^ were calculated for all estimates derived from random samples. Coverage probabilities of the 95% confidence interval for prevalent screening rates were confirmed via Monte-Carlo simulation using 10,000 replicates.

### Manual Chart Review

Institutional Review Board approval for record re-identification was requested and obtained (#18-25166). We performed a stratified random sample of charts (50 classified positive, 150 classified negative). Chart annotation criteria were serially developed and agreed upon by all reviewers after each completing a test set of 10 charts independent of the above set. Charts were annotated by the reasons for screening or the lack thereof where appropriate (see Supplemental Methods). Clinician documentation of a history of prior screening outside the institution was counted as evidence of screening. Charts were each independently reviewed and annotated by one internist and one gastroenterologist each, with all disagreements discussed and resolved. In scenarios where screening appeared to have not been performed due to a misunderstanding of the proper screening or surveillance interval, direct communication was made with the primary care provider.

## Results

The population of 50-74 year old patients with EHR data at our institution consisted of 291,420 patients, nearly a third of the total database population (See Figure 1 and Table 1). Within this cohort we identified a sub-cohort of 4,611 average risk patients empaneled in the primary care or gastroenterology clinics in the 2016-2017 period. 99% of these patients met the inclusion criteria on the basis of primary care visits within the study period. Nearly 60% of the cohort was female with an average age of 62. The racial makeup of this cohort was 42% White, 28% Asian, and 11% Black. 88% were of non-Hispanic or Latino ethnicity, and 87% declared a primary language of English. Insurance coverage was collected but as much as 95% of this information was missing.

**Table 1:** Demographics of primary care cohort at average-risk for colorectal cancer at UCSF. Other, unknown and unspecified values were excluded.

|  | *Average Risk* |  |  |
| --- | --- | --- | --- |
|  | *Primary Care Cohort* | *Screened Cohort* | *Unscreened Cohort* |
| N (%) | 4,611 | 2,835 (61) | 1,776 (39) |
| Age (years; mean ± SD) | 62 ± 7 | 62 ± 7 | 63 ± 6 |
| **Sex N (%)** |  |  |  |
| Male | 1,877 (41) | 1,127 (60) | 750 (40) |
| Female | 2,734 (59) | 1,708 (63) | 1,026 (38) |
| **Ethnicity N (%)** |  |  |  |
| Hispanic or Latino | 415 (9) | 274 (66) | 141 (34) |
| Not Hispanic or Latino | 4,076 (88) | 2,499 (61) | 1,577 (39) |
| **Race N (%)** |  |  |  |
| Asian | 1,278 (28) | 837 (66) | 441 (35) |
| Black or African American | 524 (11) | 340 (65) | 184 (35) |
| White or Caucasian | 1,949 (42) | 1,112 (57) | 837 (43) |
| **Preferred Language N (%)** |  |  |  |
| English | 4,015 (87) | 2,428 (61) | 1,587 (40) |
| Spanish | 70 (2) | 49 (70) | 21 (30) |
| Chinese – Cantonese | 176 (4) | 108 (61) | 68 (39) |
| Russian | 19 (0) | 16 (84) | 3 (16) |
| Chinese – Mandarin | 84 (2) | 57 (68) | 27 (32) |

We classified these patients by screened status based on the presence of antecedent procedure codes (see Methods) and calculated a screening rate of 61%.

We then performed manual review of 150 medical records lacking evidence of timely screening in the structured database (See Table 2). 31 patients were correctly classified as unscreened, corresponding to a negative predictive value of 21% (95% CI 15-28%). Within this group, 15 patients (48%, 95% CI 32-65%) had exams that were ordered but were not completed (eight with colonoscopy, six with FIT, one with CT Colonography). Nine unscreened patients (29%, 95% CI 16-47%) either lacked documentation for unscreened status or were incorrectly documented by the responsible physician (e.g. misunderstanding of the screening interval). Four patients (13%, 95% CI 5-29%) declined screening, and there was insufficient time to discuss cancer screening in the case of three patients (10%, 95% CI 3-26%).

**Table 2:** Reasons for True and False Classifications Identified by Manual Chart Review. The second column lists the number of charts and associated percentage of the group with 95% confidence intervals

|  |  |
| --- | --- |
| **Reasons for True Negative Classification** | **N=31, 21% (15-28%)** |
| Exams ordered but not completed | 15, 48% (32-65%) |
| --- Colonoscopy ordered but not completed | --- 8 |
| --- Fecal Immunochemical Test ordered but not completed | --- 6 |
| --- Computed Tomography - Colonography ordered but not completed | --- 1 |
| Lack of documentation or incorrect documentation | 9, 29% (16-47%) |
| Declined screening | 4, 13% (5-29%) |
| Insufficient time to discuss | 3, 10% (3-26%) |
| **Reasons for False Negative Misclassification** | **N=104, 69% (62-76%)** |
| Screening outside of UCSF | 53, 51% (41-60%) |
| Screening prior to Epic EHR implementation | 29, 28% (20-37%) |
| Database and query errors | 22, 21% (14-30%) |
| **Misclassified as Eligible for Screening** | **N=15, 10% (6-16%)** |
| Poor life expectancy, or risks outweighing benefits | 8, 53% (30-75%) |
| Above risk (personal or family history of polyps) | 6, 40% (20-64%) |
| Not primary-care empaneled | 1, 7% (0-32%) |
| **Reasons for False Positive Misclassification** | **N=2, 4% (0-14%)** |
| Ordered but incomplete Fecal Immunochemical Test | 1, 50% (9-91%) |
| Performed colonoscopy revealed inadequate bowel preparation | 1, 50% (9-91%) |

A total of 104 (69%, 95% CI 62-76%) patients had positive evidence of screening upon manual chart review. Fifty-one percent underwent screening outside of our institution (95% CI 41-60%), and 28% (95% CI 20-37%) had screening exams prior to the implementation of the Epic EHR in June 2012. The remaining 22 false negative records (21%, 95% CI 14-30%) were associated with screening exams otherwise expected to occur within the theoretical scope of the database. Some of these errors were related to screening exams performed around the time of database creation (March 2018) or *Epic* software installation (June 2012). The reasons identified for other errors were multifactorial but include errors of database creation and structure and at the level of querying.

The remaining 15 patients (10%, 95% CI 6-16%) corresponded to patients who were not actually eligible for screening. Eight patients had a poor life expectancy or were felt to have risks that outweighed the benefits of screening. Six patients were classified as above average risk on the basis of either a personal or family history of polyps. One patient was not clearly empaneled in the primary care clinic despite confirmation of two primary care office visits during the study period.

Lastly, manual review of 50 records suggested to be up-to-date with screening indicated a positive predictive value of 96% (95% CI 86-100%) (See Table 2). Two patients (4%, 95% CI 0-14%) were unscreened – one ordered but incomplete FIT and one with a prior colonoscopy but inadequate bowel preparation.

Using the aforementioned positive and negative predictive values, we calculated a corrected period prevalent screening rate of 85% (81-90%). The most common screening modality used was colonoscopy. Other notable global findings include four charts with incorrectly documented surveillance intervals. For example, one chart with a negative FIT in 2014 was incorrectly flagged for follow-up screening in 2024. We identified one patient (2%, 95% CI 0-11%) who screened positive by FIT and was referred for colonoscopy, but the referral expired. We noted occasional discrepancies between surveillance intervals proposed by the gastroenterologist and primary care physician (e.g. 5-versus 10-year follow-up).

## Discussion

Medical billing data ostensibly contain most of the fields needed to estimate screening rates and identify unscreened patients: age, procedures, diagnoses, and dates. Billing data generated in EHR during clinical operations are electronically transmitted to healthcare insurers, and therefore comprise a common substrate with their claims data – a reasonable proxy for services actually rendered. As such, one might think that these data are readily suitable to answer any of a variety of population-health questions including cancer screening rates.

A common practice for study validation involves the comparison of estimates derived from one data source to another independent and/or gold-standard dataset. However, a naïve estimation of the CRC screening rate using this database alone and without record review would have precisely matched the CDC’s estimate of 60% screened. Thus, any study of administrative data that relies on external validation due to the impossibility of internal validation (e.g. payor claims data) is not immune to the possibility of significant systematic bias.

Although this study suggests that the “out of the box” use of administrative healthcare data has the potential to be quite error prone, we also believe that it also has tremendous potential. The key to achieve this are methods capable of correcting the inherent biases to better match the data quality typical of prospectively collected, research-grade clinical data. Here we demonstrate a low-tech, interpretable, and reliable method of achieving the same end: a hybrid approach that combines clinical informatics with targeted review. More complex approaches such as natural language processing and machine learning might eventually be able to perform this task in a scalable way.

How do our results compare with previously published estimates? To our knowledge only one study from Petrik et al.^13^ has directly reported the positive and negative predictive value of EHR billing codes at identifying screened and unscreened patients. They too reported a high accuracy of identifying screened patients. By contrast, 88% (85-91%) of the patients identified as unscreened in their study were confirmed by manual chart review, compared to 21% in our study.

Several potential reasons exist for this discrepancy. For one, their study aimed to identify patients in need of screening, whereas this study aimed to accurately capture the prevalent screening rate. Our study excluded from the denominator any patient lacking a primary care relationship as well as those for whom the risks of screening outweigh the benefits. The study by Petrik et al. counted all patients with end-stage renal disease, inflammatory bowel disease, colon cancer or total colectomy towards those not needing screening. We did not informatically exclude patients with significant comorbid diagnoses or compute a Charlson Comorbidity Index; doing so would have introduced bias in our tertiary-care center where many sick patients undergo cancer screening prior to organ transplantation.

A second important difference is that our study treated as screened only those who had evidence of an adequate and complete screening exam. As such, we intentionally included a gap between the last date of study inclusion (December 2017) and the cutoff for screening completion (March 2018). Unlike the Petrik et al. study, referrals alone were not considered adequate. Although both studies accepted written documentation of a qualifying screening exam as adequate evidence, our chart review protocol included a review of the endoscopy report to confirm adequacy of bowel preparation. The two false positive cases and half of the true negative charts we identified were classified on the basis of incomplete exams.

Common reasons for misclassification of screened patients across both studies include note-based evidence of a qualifying screening exam. Half of the false negative charts we reviewed had evidence of screening elsewhere, and a quarter of the charts had evidence of screening within our institution but generated by legacy EHR software prior to June 2012. Although fully a quarter of the false negatives involved examinations performed within the expected scope of our database, some of these exams occurred at the end of 2012 or in March 2018 (the month the database was queried), suggesting errors due to incomplete data migration. However, we also noted other idiosyncratic errors at the level of database creation and querying.

A key strength of this work lies in the study methodology. This study utilized a comprehensive list of diagnosis and procedure codes developed in close collaboration with proceduralists, billing staff, and members of the quality improvement and accountable care division. Study investigators simultaneously contributed to both the query development and the chart review process, and improved both as a result. All charts independently examined by one internist and gastroenterologist each. We reported robust binomial confidence intervals and tested the coverage probability of the corrected prevalence estimate with Monte-Carlo simulation. This work was able to identify, at a fairly granular level, reasons for errors at all levels, from clinical informatics to the provision of primary care. This audit led to the identification of several clinical care errors, with clinicians informed and education provided where appropriate.

We acknowledge several limitations. First, the chart review process was challenging. Interpreting clinical notes is inherently a subjective process, and we encountered many edge cases that required discussion, criteria refinement, and imperfect resolution. The nuances of balancing of competing agenda, incorporating values and weighing risk-benefits within a time-limited clinic visit frequently do not make it to the written page. We also note other potential sources of measurement bias. We decided to accept note-based documentation as sufficient evidence that screening was performed, rather than having required the full screening report in the chart. We also suspect that relevant family history (e.g. interval diagnoses of advanced polyps) are not regularly rechecked and updated at each visit, contributing to some mismeasurement. Lastly, the specific colorectal cancer screening rates at our institution may not generalize to other primary care clinic populations.

Although billing data derived from the EHR and claims data from healthcare payors are similar, they are not identical. Claims data may capture healthcare utilization across multiple sites. EHR structured data captures local patient data irrespective of insured status or changes to insurance carrier. The EHR also carries the potential to explore a richer dataset including test results and unstructured data in the form of clinical notes. Both systems are subject to breaks and discontinuities as patients leave and enter (or re-enter), as well as the errors inherent to intrinsically complex, non-research grade data.

Our results indicate that the primary care apparatus at our institution is effective at performing CRC screening. Nevertheless, we see several potential areas of improvement. Improved documentation of the CRC screening decision and the disposition of screening referrals, regular updating of family history, and greater communication between gastroenterologists and internists will help all healthcare institutions improve their screening rates. They may also improve informatic ascertainment of screened status in combination with technologies such as natural language processing, optical character recognition, and deep learning (Table 3).

**Table 3:** Potential solutions to improve informatic classification and CRC screening.

| **Reasons for True Negative Classification** | **Potential Solutions** |
| --- | --- |
| Exams ordered but not completed |  |
| --- Colonoscopy ordered but not completed | -More transparent documentation of referral status and outcome<br>-Clinic-based patient outreach |
| --- Fecal Immunochemical Test ordered but not completed | -Clinic-based patient outreach |
| --- Computed Tomography - Colonography ordered but not completed | -More transparent documentation of referral status and outcome<br>-Clinic-based patient outreach |
| Lack of documentation or incorrect documentation | -Improved primary care education<br>-Improved Gastroenterologist-Primary Care communication |
| Declined screening | -Improved patient education |
| Insufficient time to discuss | -Clinic-based strategies to encourage follow-up |
| **Reasons for False Negative Misclassification** |  |
| Screening outside of UCSF | -Patient-approved data sharing, harmonization and interoperability<br>-Natural language processing<br>-Optical character recognition<br>-Deep learning |
| Screening prior to Epic EHR implementation | -Institutional investment in clinical data integration and harmonization |
| Database and query errors | -Recruitment, training and funding for more clinical informaticians, especially clinician-investigators<br>-Institutional investment in clinical data integration and harmonization |
| **Misclassified as Eligible for Screening** |  |
| Poor life expectancy, or risks outweighing benefits | -Deep learning with natural language processing |
| Above risk (personal or family history of polyps) | -Natural language processing<br>-Improved family history taking practices<br>-Patient consent for EHR data-sharing, chart-linkage by familial relationship |
| Not primary-care empaneled | -Deep learning with natural language processing |
| **Reasons for False Positive Misclassification** |  |
| Ordered but incomplete Fecal Immunochemical Test | -EHR flag/reminders to repeat screening |
| Performed colonoscopy revealed inadequate bowel preparation | -Natural language processing<br>-EHR flag/reminders to repeat screening |

However, the greatest challenge to the future of clinical informatics lies in the problem of bias in observational data. Identifying and managing bias is fundamentally a task that requires humility, vigilance, and the collaborative engagement of diverse stakeholders and domain experts who understand the provenance and meaning of the data. It requires that we stress test our data openly and often before drawing conclusions or taking action.

## Data Availability

Please contact corresponding author for questions regarding data availability.

## Acknowledgements

We thank Marlene Herrera and Sara Coleman-Hernandez for useful technical input. We thank Boris Oskotsky and Dana Ludwig for database creation and management. We thank members of the UCSF Gastroenterology Division as well as Biostatistics and Epidemiology Department for valuable discussion.

## Notes

**Conflicts of Interest:** None relevant to this publication

**Financial Support:** UCSF Bakar Computational Health Sciences Institute and the National Center for Advancing Translational Sciences of the National Institutes of Health under award number UL1 TR001872. VAR was supported by the National Institute of Diabetes and Digestive and Kidney Disease of the National Institutes of Health grant under award number T32 DK007007-42.

### Competing Interest Statement

The authors have declared no competing interest.

### Funding Statement

UCSF Bakar Computational Health Sciences Institute and the National Center for Advancing Translational Sciences of the National Institutes of Health under award number UL1 TR001872. VAR was supported by the National Institute of Diabetes and Digestive and Kidney Disease of the National Institutes of Health grant under award number T32 DK007007-42.

### Author Declarations

All relevant ethical guidelines have been followed and any necessary IRB and/or ethics committee approvals have been obtained.

Any clinical trials involved have been registered with an ICMJE-approved registry such as ClinicalTrials.gov and the trial ID is included in the manuscript.

